# Randomized Trial Protocol: Head-to-Head Comparison of Two Ambient Artificial Intelligence Documentation Tools on Clinician Usability

**DOI:** 10.64898/2026.11.07.26367514

**Authors:** Aaron T. Chin, Kenny Leung, Sitaram Vangala, Hawkin Woo, Thomas Kingsley, Alexander Sandhu, Laleh Jalalian, Eric M. Cheng, John N. Mafi, Paul J. Lukac

**Author notes:** Co-first authors (Aaron T. Chin, Kenny Leung). Co-senior authors (John N. Mafi, Paul J. Lukac). Trial registration: Submitted to ClinicalTrials.gov, pending NCT assignment. Protocol version: v1.0, 11/7/2026.

## Abstract

**Background:** Ambient AI documentation reduces documentation burden and electronic health record (EHR) time for ambulatory clinicians. Newer features also queue orders and offer some level of coding support for clinician review. This study compares two commercial products deployed at UCLA Health.

**Objective:** To compare Nabla and Epic Systems, Inc.’s ambient AI solution among ambulatory clinicians at UCLA Health on usability, documentation burden, perceived note quality, adoption, EHR time and productivity, clinician-reported experience with order queueing and coding support, coding and billing patterns and integrity, work productivity, and patient-reported communication experience.

**Methods:** Two-arm parallel-group randomized trial, 1:1, among 374 ambulatory clinicians at a single academic health system who hold an active license for a standalone ambient application (Nabla). Randomization is covariate-constrained where cohort size permits, balancing age, sex, half-day clinic sessions, specialty group, and baseline System Usability Scale (SUS) score. Intervention runs 11/9/2026 to 1/17/2027, with the first four weeks as lead-in. The primary outcome is SUS score at follow-up, compared between arms. Analysis is by intention-to-treat using mixed-effects models.

**Ethics and dissemination:** Determined exempt by the UCLA Institutional Review Board (IRB-26-2147). Survey completion indicates consent. No patient-level data are collected. Findings will directly inform operational decision making and will be published regardless of direction.

## Background and Rationale

Clinical documentation is a leading contributor to clinician electronic health record (EHR) time and burnout.^1^ Ambient artificial intelligence (AI) scribes, which draft a clinical note from the patient encounter, alleviate that burden: randomized trials at UW Health and at our institution, the latter comparing two vendors against usual documentation, found reductions in documentation time and burnout-associated outcomes.^2,3^ Physician engagement with AI tools has risen rapidly, with 81% of physicians reporting awareness or use of health AI in 2026, up from 66% in 2024.^4^

Expanded ambient capabilities, including order facilitation and coding support, have been anticipated as the next stage of the technology.^5^ Both tools in this trial queue orders for clinician review and offer some level of coding support. That turns the output from a draft note into billable codes and actionable orders, shifting risk toward coding accuracy, order appropriateness, and safety. It also shifts the clinician from producing clinical content to verifying it. A recent meta-analysis reported ambient documentation to be associated with reduced temporal demand, effort, and burnout, while imaging AI and decision support showed mixed or paradoxically increased workload, and attributed the difference to verification burden, the cognitive cost of reviewing and reconciling AI output with clinical judgment.^6^ Separately, ambient scribes may function as “coding intensity” engines, inviting payer downcoding and risk score recalibration.^7^ To our knowledge, no randomized study has compared the performance of two tools in a head-to-head RCT with ambient documentation bundled with order queueing and coding support.

The primary aim is to determine whether perceived usability differs between the two tools. Results of the RCT will directly inform operational decision making on the purchase of one or the other tool in the context of a learning health system. Additional aims compare efficiency, note quality, clinician-reported experience with order queueing and coding support, diagnosis capture and order activity, safety, workload, adoption, productivity, and patient-reported communication experience. Given the absence of prior head-to-head data on these functions, hypotheses are direction-agnostic.

## Methods

This protocol follows the SPIRIT 2025 and SPIRIT-AI reporting guidelines.

### Design and setting

Two-arm parallel-group randomized controlled trial, 1:1, no crossover, in ambulatory clinics at UCLA Health, which uses a single enterprise EHR (Epic Systems, Inc. [Verona, WI]). The intervention period runs 11/9/2026 to 1/17/2027.

Both tools are deployed operationally by UCLA Health as replacements for a pre-existing, partially integrated version of Nabla. Clinicians who complete the baseline survey are randomized to Nabla Ambient Voice Recognition (AVR), a fully electronic health record (EHR)-integrated version of Nabla, or Epic Chart with Art, Epic’s native ambient functionality. Clinicians who do not complete the baseline survey or opt out of the study are assigned a base version of Nabla AVR, which lacks ancillary features such as order queueing, and are operationally sequestered from the trial. Clinicians will also complete a follow-up survey at the completion of the study period, with results reported to leadership in aggregate only.

### Participants

Ambulatory physicians and advanced practice providers holding an active Nabla license at solicitation. Recruitment is achieved by email solicitation, while enrollment is by survey completion.

To be included in the study, clinicians must practice at an outpatient UCLA Health Clinic(s); have used the prior version of Nabla within the preceding 90 days; and complete a baseline survey.

Physicians in training and clinicians with fewer than one half-day clinic session per week are excluded.

### Randomization

Allocation is 1:1 among clinicians who complete the baseline survey. Covariate-constrained randomization will be used to balance arms on age, sex, half-day clinic sessions per week, specialty group, and baseline System Usability Scale score. The analysis adjusts for these covariates in either case, so the analytic approach does not depend on the allocation method.

The enrolled cohort is allocated in a single step after the baseline survey closes. A designated honest broker holds the key linking clinician identity to study identifier, so the study team does not know which clinicians are in each arm. Blinding for the users is not possible, as the intervention is the documentation interface.

### Intervention

#### Tools and versions

Both arms use vendor-developed ambient documentation tools that apply generative AI to encounter audio. The underlying models are proprietary. Each vendor will be asked to provide the model and software versions deployed during the intervention period, and versions and the dates of any updates will be recorded. If a version update occurs during the trial, the prespecified analysis is unchanged.

#### Scope

Neither tool draws on prior notes or other chart content. Input is limited to the encounter transcription. A separate Epic AI chart summarization feature is available to all participants in both arms. It is not part of either intervention.

#### Integration

Nabla AVR is integrated into the Epic EHR. Epic Chart with Art is native Epic functionality.

#### Input data

The input to both tools is encounter transcription. In each encounter, the clinician obtains verbal patient consent and then starts the recording. Audio recordings are not retained.

#### Eligible encounters

Tools may be used in any ambulatory encounter, including encounters conducted in a language other than English and encounters using an interpreter. Use in non-English encounters is limited to clinicians certified in that language.

#### Outputs

Both tools produce a draft clinical note and queued orders. Epic Chart with Art additionally produces queued diagnoses. Nabla AVR additionally produces a hierarchical condition category (HCC) documentation check and, for selected specialties, an evaluation and management (E/M) level recommendation (Table 1).

**Table 1.** Features by Arm.

| **Feature** | **Nabla AVR** | **Epic Chart with Art** |
|---|---|---|
| Ambient note generation | All participants | All participants |
| Order queueing | All participants | All participants |
| Diagnosis queueing | Not available | All participants |
| HCC documentation check | On by default for primary care (family medicine, internal and general medicine, pediatrics); off by default for other specialties, with the option to turn it on | Not available |
| E/M level recommendation | Family medicine, internal and general medicine, pediatrics, psychiatry, cardiology, neurology | Not available |

#### Use of outputs

All generated content requires clinician review. In this pragmatic trial, every generative AI output, including drafted notes, order queues, etc., is manually reviewed by a clinician as per routine workflows and quality assurance. Notes are edited and signed by the clinician, and queued orders and diagnoses take effect only when the clinician accepts them. The HCC documentation check and E/M level recommendation are advisory.

#### Access

Both tools are proprietary commercial products. The models and code are not accessible to the investigators or available for reuse.

The first four weeks (11/9/2026 to 12/6/2026) are a lead-in to account for the natural learning curve, while the remaining six weeks (12/7/2026 to 1/17/2027) constitute the comparison period. Use is optional, and all participants receive training on tool functionality and the limitations of generative AI output. Training combines vendor-prepared materials and virtual support sessions and is equivalent across arms.

The enterprise default ambient documentation tool is the base version of Nabla AVR, which provides note- writing functionality only. Clinicians who discontinue their assigned tool will revert to this default and remain in their randomized arm for analysis.

### Comparator

Each arm is the comparator for the other. Because the trial’s primary purpose is operational and quality improvement, the head-to-head design compares the latest version of each tool at the time of the trial. Both are observed over the identical calendar window and transition from the same retiring application, so seasonal effects, secular trends, and learning curves are shared. Because feature sets differ, arms are compared as deployed. There is no usual control group.

### Primary outcome

System Usability Scale at follow-up, compared between the two arms.^8^ This was chosen first because of its widespread use in technology assessment research and second because it fits the primary objective of this trial, which is to inform health system operational decision making on which tool to purchase. This survey instrument includes ten items, each on a five-point Likert scale, with individual results reported on a scale of 0 to 100. Wording is verbatim with the assigned tool substituted for “the system.” The SUS is also administered at baseline referencing the standalone application (Nabla) and is included in the model as a baseline measurement.

### Exploratory outcomes: clinician survey

Baseline survey items referencing a documentation tool refer to the retired standalone application, and the corresponding follow-up items refer to the assigned tool. This provides a within-clinician baseline for note quality, documentation experience, and task load. Burnout is measured independent of tool. Items on order queueing and coding support are administered at follow-up only, since the standalone application did not offer this functionality, and are displayed only to participants who report using the feature.

| Outcome | Timing | Description |
|---|---|---|
| Net Promoter Score | Baseline and follow-up | Likelihood of recommending the tool to a colleague, 0 to 10. NPS is the percentage of promoters (9 to 10) minus the percentage of detractors (0 to 6), range -100 to 100, calculated by arm.<sup>9</sup> |
| Documentation experience | Baseline and follow-up | Four items on a 5-point agreement scale: time saved, note style fit, editing burden, trust in the tool. Each reported separately. Investigator-developed. |
| Perceived note quality | Baseline and follow-up | Four PDSQI-9-derived attributes on a 5-point frequency scale: accurate, thorough, organized, succinct. Each reported separately.<sup>10</sup> |
| Physician task load | Baseline and follow-up | Four NASA-TLX items referencing closing out visits in the EHR: mental demand, temporal demand, effort, physical demand. Higher scores indicate greater task load.<sup>11</sup> |
| Burnout | Baseline and follow-up | Mini-Z 2.0 single item, five options. Reported as the percentage of participants meeting the standardized dichotomous definition of burnout.<sup>12</sup> |
| Diagnosis queueing | Follow-up | Epic Chart with Art arm. Six items on a 5-point frequency scale: use, time saved, completeness, wrong diagnosis, exaggerated severity or specificity, and unsupported suggestions that would have been inappropriate to accept without review. Response distributions. |
| HCC documentation check | Follow-up | Nabla AVR arm. Three items on a 5-point frequency scale: documentation changes prompted by the check, MEAT (Monitor, Evaluate, Assess, & Treat) criteria flagged as missing when addressed in the encounter, and time saved. Response distributions. |
| E/M level recommendation | Follow-up | Nabla AVR arm. Three items on a 5-point frequency scale: E/M level changed after review, recommendation higher than documentation supported, and recommendation lower than documentation supported. Response distributions. |
| Order queueing | Follow-up | Six items on a 5-point frequency scale: use, time saved, completeness, wrong order, wrong order details, and orders that would have been clinically inappropriate if placed without review. Response distributions by arm among participants reporting use. |
| Reported safety | Follow-up | Screener for safety concerns attributable to the tool, with severity (near miss; reached patient without harm; resulted in harm), institutional filing, and description. Number of participants reporting a concern by arm and severity. Investigator-developed; no validated instrument exists for this construct. |
| Forced choice | Follow-up | Preference for the assigned tool versus the retired standalone application, with rationale. |
| Free-text feedback | Follow-up | Open responses on note inaccuracies, omissions, and other concerns or unintended effects, analyzed by thematic analysis. |

### Exploratory outcomes: operational data

Baseline is the six-month retrospective period from 5/2/2026 to 11/1/2026. The four-week lead-in (11/9/2026 to 12/6/2026) is excluded from both baseline and the comparison period. Operational outcomes are analyzed as change from baseline, and arms are compared on that change. Findings from exploratory operational outcomes may be reported in separate, subsequent publications rather than included in the initial primary publication.

| Outcome | Baseline | Description |
|---|---|---|
| EHR time | 6 months, per clinician | Epic Signal, in minutes: time in notes per note, time in orders per appointment, time outside scheduled hours per scheduled day. Pajama time reported descriptively as a component. |
| Tool utilization | Standalone application usage, up to 6 months prior | Proportion of eligible ambulatory encounters documented with the assigned tool. |
| Diagnosis capture* | 6 months, per clinician | Total final diagnosis codes entered per encounter. |
| Diagnosis capture integrity | Follow-up, per clinician | Coding operations reviews a sample of comparison-period encounters per arm for code support against the note; the review has no baseline. Performed as per routine operational workflows; only aggregate results reach the study team. |
| Order activity* | 6 months, per clinician | Orders per encounter, cancellations per encounter, and duplicate order alerts per encounter. The latter two are proxies for medical error. |
| Productivity | 6 months, per clinician | Work relative value unit (wRVU) per clinical session. |
| Billing | 6 months, per clinician | Proportion of encounters at each E/M service level.<br>Aggregate standardized wRVUs. |
| Patient experience | 6 months, per clinician | Clinician & Group Consumer Assessment of Healthcare Providers and Systems (CG-CAHPS) provider communication composite, aggregated per clinician for baseline and the comparison period and compared between arms.<sup>13</sup> |

*Descriptive. The tool only suggests; what is recorded is what the clinician enters, so codes and orders cannot be attributed to the tool.

### Subgroup analyses

Prespecified and exploratory, all at the clinician level: years since training; specialty group; baseline time in notes per encounter; baseline clinic volume; age group; sex.

### Data collection and sources

Baseline data are collected by Qualtrics survey 10/11/2026 to 10/28/2026, before deployment. Follow-up data are collected 1/18/2027 to 2/14/2027 and include survey responses and EHR metrics from Epic Signal and Caboodle, compared against a six-month retrospective baseline.

Surveys are administered by the research team through Qualtrics. A designated honest broker within health system operations, who is not a member of the study team, links survey responses to EHR metrics and treatment assignment under a study identifier. The link between identity and study identifier is held solely by the honest broker and destroyed at study closure.

No patient-level protected health information is collected. The diagnosis coding audit and the CG-CAHPS aggregation are routinely performed within health system operations. Coding audit results are released to the study team at the arm level and CG-CAHPS results at the clinician level, both de-identified. EHR-derived and patient experience data are de-identified, aggregated, and analyzed at the clinician level. Age is released in bands, and specialty groups with fewer than ten clinicians are collapsed.

### Statistical analysis

Analysis follows intention-to-treat; all randomized participants are analyzed in their assigned arm regardless of use. The primary estimand is the difference between arms in mean System Usability Scale score at follow- up, interpreted as the effect of switching to Nabla AVR versus Epic Chart with Art on usability. The primary analysis uses a linear mixed model with clinician random effects, with baseline and follow-up scores as repeated measures, fixed effects for arm, period, and their interaction, and adjustment for sex, age, specialty group, and half-day sessions. Modeling baseline as a timepoint accounts for baseline usability within the model.

Other survey outcomes measured at baseline and follow-up use the same specification, and EHR metrics use the retrospective baseline, lead-in, and comparison period as periods. Likert-type items are analyzed with linear mixed models. Other outcome types use generalized linear mixed models with a similar specification (e.g., mixed-effects logistic regression for binary outcomes). This rapid cycle RCT makes a mid trial version update or upgrade less likely. However, in the event of an unanticipated version upgrade, we will proceed with the original pre-specified analysis plan of comparing outcomes in the last 6 weeks of the trial, to give a pragmatic, overall effect of deploying dynamic AI tools in real-world settings.

The intention-to-treat analysis of survey outcomes is robust to dropout under a missing at random assumption. Baseline characteristics of participants who complete the follow-up survey are compared descriptively with those who do not, overall and by arm. The primary outcome uses a 0.05 significance level; other analyses report estimates and 95% confidence intervals without hypothesis testing. Subgroup analyses add an arm by subgroup interaction term and are not powered to detect interaction. Clinicians who withdraw after learning their assignment will be analyzed as randomized. If more than 10% of clinicians withdraw from either group, an instrumental variables analysis will be performed to estimate the complier average causal effect and correct for the resulting dilution of the intention-to-treat estimate. Analysis will be performed using R version 4.6.1.

### Sample size

A total of 374 clinicians enrolled. This provides >80% power to detect differences as small as 0.32 standard deviations at a two-sided alpha of 0.05. The detectable difference reflects actual enrollment rather than a minimally important difference.

### Data monitoring

No data monitoring committee and no stopping rules. The trial is deemed to present minimal risk, both tools are deployed operationally regardless of participation, and all generated content requires clinician review before a note is signed or an order placed. Affirmative safety responses are reviewed by the study team and referred through institutional patient safety channels where appropriate.

### Withdrawal and data access

Clinicians who wish to discontinue their assigned tool are encouraged to contact the support team first, and the reason for discontinuation will be recorded. Clinicians who discontinue revert to the enterprise base version of Nabla AVR and remain in their randomized arm for analysis.

### Patient involvement

Patients were not involved in the design, conduct, reporting, or dissemination of this research. Outcomes are analyzed at the clinician level, including patient-reported communication measures aggregated to the clinician.

### Ethics and consent

Determined exempt by the UCLA Institutional Review Board (IRB-26-2147). Clinicians are recruited by email. The baseline survey opens with a study information sheet. Survey completion indicates informed consent and is required for randomization. Nonparticipants receive a replacement tool operationally, and the standalone application is retired regardless of participation. Individual responses are not shared with supervisors or used for employment, credentialing, productivity, or performance evaluation. Results are reported in aggregate.

### Privacy and data security

Data are stored on secure institutional systems with access restricted to study personnel. The quantitative dataset is limited to clinician-level survey responses and aggregated clinician-level metrics, contains no clinician or patient identifiers, and is not readily re-identifiable by the study team. The code key is held solely by the honest broker and destroyed at study closure.

## Discussion

To our knowledge, this is the first head-to-head randomized trial comparing two ambient AI documentation tools with distinct embedded features that support clinical workflows beyond note-writing, including order queueing and coding support. These features introduce more AI-generated content that clinicians may need to verify, which can add cognitive burden.^6^ A real-world, current-state comparison between the two tools will assess how perceived note quality and feature usefulness affect user-reported usability, task load, and burnout.^8,11,12^ This study will also provide insight into how these features may affect time efficiency, diagnosis capture, and work productivity.

This study design has several strengths. Clinicians are randomized in a health system serving a diverse patient population. The study also evaluates the tools in their current states, without matching feature sets. This makes outcomes harder to attribute to any single feature, but it is a pragmatic approach, since competing tools are rarely identical. It also allows the study to inform operational decisions about which feature offerings are most desirable to clinicians, which was the primary purpose of this operational study.

There are several limitations to this single-center study. All participants used Nabla before randomization, so clinicians assigned to Chart with Art switch tools while those assigned to Nabla stay on a familiar platform with added features. Participants may also discontinue their assigned tool and revert to the base version of Nabla AVR, which is functionally similar to the legacy application, and this may increase discontinuation and dilute differences between arms. The short intervention of 10 weeks, including a 4-week lead-in, may not capture the full adoption curve. However, as AI tools evolve rapidly, a longer study period may risk assessing an outdated version of the tool. The trial is conducted at a single academic health system, which may limit generalizability.

## Declarations

### Trial registration

Submitted to ClinicalTrials.gov, pending NCT assignment.

### Trial status

Recruitment closed on 10/28/2026 with 374 clinicians enrolled. The intervention period begins on 11/9/2026.

### Data availability

Datasets will not be publicly available due to institutional restrictions. De-identified, aggregated data may be available from the corresponding author upon reasonable request and subject to institutional approvals.

### Ethics

Determined exempt by the UCLA Institutional Review Board (IRB-26-2147). No patient-level data are collected.

## References

1. Arndt BG, Beasley JW, Watkinson MD, et al. Tethered to the EHR: Primary Care Physician Workload Assessment Using EHR Event Log Data and Time-Motion Observations. Ann Fam Med 2017;15:419–426. DOI: 10.1370/afm.2121.

2. Afshar M, Baumann MR, Resnik F, et al. A Pragmatic Randomized Controlled Trial of Ambient Artificial Intelligence to Improve Health Practitioner Well-Being. NEJM AI 2025;2. DOI: 10.1056/aioa2500945.

3. Lukac PJ, Turner W, Vangala S, et al. Ambient AI Scribes in Clinical Practice: A Randomized Trial. NEJM AI 2025;2. DOI: 10.1056/aioa2501000.

4. American Medical A. 2026 Physician Survey on Augmented Intelligence. Chicago, IL: AMA Center for Digital Health and AI, 2026/03 2026. (https://www.ama-assn.org/system/files/physician-ai-sentiment-report.pdf).

5. Ohde JW, Thompson A, Liu Z, et al. Barriers and opportunities of scaling ambient AI scribes for clinical documentation across diverse healthcare settings. NPJ Digit Med 2026;9:369. DOI: 10.1038/s41746-026-02554-0.

6. Gong EJ, Bang CS, Lee JJ. Cognitive Workload and Mental Burden in Health Care Professionals Interacting With AI: Systematic Review and Meta-Analysis. J Med Internet Res 2026;28:e93618. DOI: 10.2196/93618.

7. Dai T, Kvedar JC, Polsky D. Policy brief: ambient AI scribes and the coding arms race. NPJ Digit Med 2025;8:780. DOI: 10.1038/s41746-025-02272-z.

8. Lewis JR. The system usability scale: past, present, and future. International Journal of Human–Computer Interaction 2018;34:577–590.

9. Reichheld FF. The one number you need to grow. Harv Bus Rev 2003;81:46–54, 124. (https://www.ncbi.nlm.nih.gov/pubmed/14712543).

10. Croxford E, Gao Y, Pellegrino N, et al. Development and validation of the provider documentation summarization quality instrument for large language models. J Am Med Inform Assoc 2025;32:1050–1060. DOI: 10.1093/jamia/ocaf068.

11. Melnick ER, Harry E, Sinsky CA, et al. Perceived Electronic Health Record Usability as a Predictor of Task Load and Burnout Among US Physicians: Mediation Analysis. J Med Internet Res 2020;22:e23382. DOI: 10.2196/23382.

12. Linzer M, McLoughlin C, Poplau S, et al. The Mini Z Worklife and Burnout Reduction Instrument: Psychometrics and Clinical Implications. J Gen Intern Med 2022;37:2876–2878. DOI: 10.1007/s11606-021-07278-3.

13. Agency for Healthcare R, Quality. CAHPS Clinician & Group Survey. 2026 (https://www.ahrq.gov/cahps/surveys-guidance/cg/index.html).

